# An Innovative Visual Approach to Monitor Simultaneously Two Dimensions of Progress in Longevity: An Application to French and German Regions

**DOI:** 10.1101/2023.09.13.23295507

**Authors:** Florian Bonnet, Sebastian Klüsener, France Meslé, Michael Mühlichen, Pavel Grigoriev

**Affiliations:** French Institute for Demographic Studies (INED), Aubervilliers, France; Federal Institute for Population Research (BiB), Wiesbaden, Germany; University of Cologne, Cologne, Germany; Vytautas Magnus University, Vilnius, Lithuania

## Abstract

**Background:** Both enhancing life expectancy as well as diminishing inequalities in lifespan among social groups represent significant goals for public policy. However, there is a lack of methodological tools to simultaneously monitor progress in both dimensions. Additionally, there is a consensus that absolute and relative inequalities in lifespan must be scrutinized together.

**Methods:** We introduce a novel graphical representation that combines national mortality rates with social inequalities, considering both absolute and relative measures. We use French and German data stratified by place of residence to illustrate this representation.

**Results:** For all-age mortality we detect for France a rather continuous pace of decline in both mortality levels and variation. In Germany, substantial progress was made in the 1990s, which was mostly driven by convergence between eastern and western Germany, followed by a period with less progress. Age-specific analyses reveal for Germany some worrying regional divergence trends at ages 35-74 in recent years. This is particularly pronounced among women.

**Conclusion:** Our novel visual approach allows evaluating easily the dynamics of societal progress in terms of longevity, and facilitates meaningful comparisons between populations, even when their current mortality rates differ. The methods we employ can be reproduced easily in any country with longitudinal mortality data stratified by relevant socio-economic information or regions. It is both useful for scientific analyses as well as policy advice.

**Key messages:** *What is already known on this topic:* Improving life expectancy as well as reducing social inequalities in longevity are major public policy objectives. However, there is a lack of proper methodological tools to evaluate progress on these objectives.

*What this study adds:* This study proposes an innovative graphical representation that combines national mortality and social inequalities in both absolute and relative terms in order to assess the dynamics of societal progress in longevity and make relevant comparisons between populations whose mortality rates are not at the same level nowadays.

*How this study might affect research, practice or policy:* Methods are freely and easily reproducible for all countries with longitudinal mortality data stratified by socio-economic information or geographic regions.

## Introduction

Living conditions and survival chances have strikingly improved since the beginning of the 20th century in a large number of countries. Yet, this improvement does not necessarily indicate societal progress in human development, as such progress should benefit equally to all population strata. Consequently, policy makers and scholars are increasingly interested in the issue of inequality.

Income inequality has long been a significant but controversial topic in economics. Some scholars have expressed concerns that resources would become increasingly concentrated over time, while others believed that market forces would reduce inequality after an initial upward trend in the early stages of development [1]. The lack of comprehensive statistical indicators describing long-term trends at the individual level has hindered the resolution of this debate. Nevertheless, recent influential publications [2, 3, 4] highlight a substantial decline in income inequality during the first half of the 20th century, followed by a recent increase, particularly substantial in the United States [5, 6].

In longevity research, recent studies have demonstrated how assessing simultaneously both mean and dispersion measures of lifespan is important to study national populations. First, such a framework allows better international comparisons [7, 8, 9, 10] by pinpointing countries with high lifespan dispersion after controlling for mean values. For instance, France and the United States reached male life expectancy of 75 years with a higher level of lifespan inequality than Sweden or England and Wales did. Second, it is now not sufficient to study the trends in lifespan mean value to know the trends in lifespan inequality since the strength of their correlation is waning over time. Historically, the joint improvement was due to the mortality decline at young ages [11], driven by reduction in both infectious and childbearing mortality [12]. Nowadays, this joint improvement is no longer evident [13, 14, 15]: the reduction of old-age mortality is the main source of life expectancy improvement, while infant and maternal mortality have negligible impact. Moreover, mortality at working ages plays a modest role in this improvement, or in some cases (like the US) even works in the opposite direction [16].

Focusing on national population stratified by socioeconomic status (such as education level [17], socio-professional category [18], or income [19]) or by place of residence [20, 21, 22, 23, 24], other studies usually solely focus on the evolution of lifespan inequality, using a large panel of indicators expressed alternatively in absolute or relative terms. However, making conclusions about the dynamics of societal progress through these inequality measures alone is complex and requires some caution. Even though absolute lifespan inequality decreases when national lifespan increases [25] over the long run, a decrease in lifespan inequality might come along with a decrease in national lifespan: [23] observed a few periods of “*downward convergence*” between 1800 and 1880 in France, when epidemics highly afflicted vanguard regions; the 2003 heatwave is another recent example. Moreover, [26, 27] suggest that there is a heuristic rule linking very low overall mortality rates and high relative inequality (“When two groups differ in their susceptibility to an outcome, the rarer the outcome, the greater the disparity in experiencing the outcome and the smaller the disparity in avoiding the outcome.”). This could explain why inequality is higher in Nordic countries than other European countries, despite their lower national mortality rates [28, 29].

In this paper, we consider these limits and propose a new framework to assess the societal progress in longevity. Our main contribution is an innovative graphical representation that combines nationwide mortality and inequality in both absolute and relative terms. This way, we reveal in one graphic how societal progress in mortality evolves over time, and disentangle the periods of absolute convergence/divergence, relative convergence/divergence and improvement/worsening of national mortality. Moreover, we strongly advocate that this representation allows better comparisons by sex or across countries since the differences in nationwide mortality observed partly explain the greater or lesser absolute inequality.

## Data and Methods

To illustrate our new approach, we rely on long-term mortality series for French and German regions. French death and population counts were obtained from the French Human Mortality Database [30] on the level of 95 *départements* of metropolitan France (NUTS-3, the finest geographical level of the classification used by Eurostat), for the period from 1970 to 2019 and by quinquennial age groups (0, 1–4, 5–9, …, 90–94, 95+). For Germany, we obtained death and population counts from the German federal and state statistical offices and harmonized them on the level of 96 *Raumordnungsregionen* for the period from 1992 to 2019. Deaths and population counts are available for the same ages except for the broader groups 1–14 and 90+.

We computed standardized death rates (SDRs) for the whole population as well as for broad age groups (0, 1–14, 15–34, 35–64, 65–74, 75+) using the European Standard Population 2013. To reduce the regional volatility, we pooled death and population counts by 3-year periods. In this way, 1995 refers to the period 1994–1996. Only 2019 refers to the 2-year period 2018– 2019.

To assess the achievement of what we call “societal progress in mortality”, we considered two sets of indicators. National SDR informs to what extent the mortality improves (or worsens) at a national level (First objective). Applied on regional SDRs, the standard deviation – as an absolute measure – and the coefficient of variation – as a relative measure – inform to what extent this improvement is similar across the considered regions (second objective). We computed both weighted and non-weighted inequality measures.

All calculations are performed in R version 4.0.3 [31].

## Results

Figure 1 presents the pathways of societal progress in mortality for France (blue) and Germany (orange), by differentiating men (lighter colors) and women (darker colors). Each point describes the couple of values of the national SDR (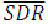) and the standard deviation of regional SDRs (*SD*_SDR_) for a 3-year period. Points are more transparent and smaller the further away they are back in time.

**Figure 1.**
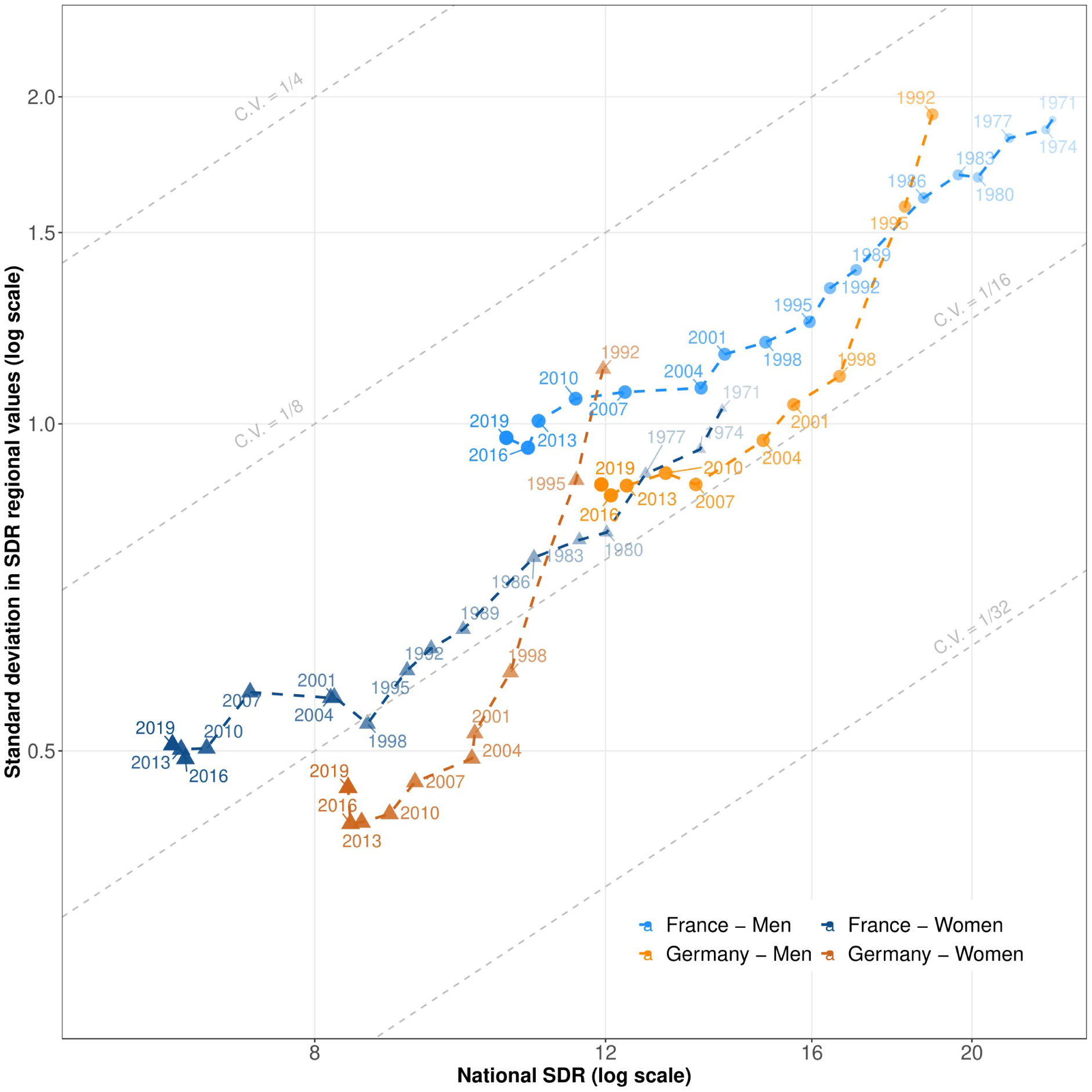
Pathways of national SDRs and regional inequalities in France (1970–2019) and Germany (1992–2019)

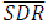 and *SD*_SDR_ are plotted in a logarithmic scale to evaluate the relative variation of these values between two periods. Importantly, one can see the relative variation of the coefficient of variation (*CV*_SDR_) using the following relationships:

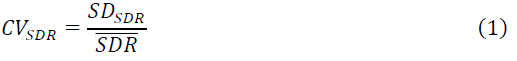

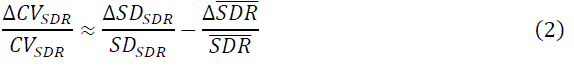

When the relative variation of *SD*_SDR_ is higher than the relative variation of 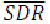, *CV*_SDR_ increases; graphically, the pathway goes to an upper grey line, which delineate values for coefficients of variation. On the opposite, when the relative variation of *SD*_SDR_ is lower than the relative variation of 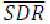, *CV*_SDR_ decreases; graphically, the pathway goes to a lower grey line.

Figure 1 reveals huge societal progress in mortality for French women between 1971 and 1998: the dramatic fall in female mortality (39%) came along with an even more rapid fall in absolute regional inequalities (49%). This difference in decline paces led to a decline in relative regional inequalities: the coefficient of variation decreased by 16% and reached the value of 1/16. Between 1998 and 2007, female mortality continued to decline (except between 2001 and 2004 because of the 2003 heatwave [32, 33]), but this progress did not come along with decreasing regional inequalities: a period of absolute and relative divergence replaced the strong convergence of previous years. Since 2010, societal progress in mortality is low: both nationwide mortality and regional mortality inequalities have barely declined. The male pathway shows strong similarities with the female one, except for the recent period: absolute regional inequalities continued to decrease between 1998 and 2007, as did nationwide mortality between 2010 and 2019.

The German case is quite different from France. In 1992, just after German reunification, regional inequalities were much higher for the same mortality level observed during the early 1980s in France. Societal progress in mortality was impressive for men from 1992 to 1998 (1992 to 2004 for women): the sharp fall in mortality (–12%) was mirrored by an even greater fall in absolute regional inequalities (–47%), leading to a consequent fall in relative regional inequalities (–44%). Thereafter, societal progress in mortality continued, but the decline in absolute regional inequality and in national mortality occurred at a similar pace, leading to almost no variation in relative regional inequalities. Importantly, this pace of societal progress in mortality has decreased and is almost nil since 2013 for both men and women. In addition, absolute inequalities for women highly increased from 2016 to 2019.

Figure 1 also enables both comparisons between the sexes and across countries. Pathways were more favourable for women than for men in France: male mortality in 2007 was equivalent to female mortality in 1980, but absolute regional inequalities were 20% higher. In Germany, pathways were also more favourable for women than for men during recent years. Finally, pathways were more favourable in Germany than in France during the period covered by our data, except for the early and mid-1990s.

The strong convergence between western and eastern Germany largely explains the strong societal progress in mortality experienced by Germany during the 1990s. This is illustrated by Figure A1 in the online supplementary Appendix A, which compares the pathways for Germany as a whole (orange) to those for western (violet) and eastern Germany (green). It reveals how much higher mortality rates were in the east than in the west in 1992 (+16% for men, +15% for women). The strong convergence between the SDRs of the two parts of the country largely explains the remarkable convergence observed in Germany between 1992 and 2004. Moreover, relative regional inequalities largely decreased during this period in eastern Germany, which reached the more favourable pathway of western Germany. Since 2004, the decrease of mortality came along with constant or increasing absolute regional inequalities in the two parts, which resulted in a rapid increase of relative regional inequalities. Finally, regional inequalities are now higher in the west than in the east as compared to the early 1990s.

We go further in our analysis by disentangling SDRs by age groups. Figure 2 replicates Figure 1 for six age groups in France. First, it shows once again that the decline in mortality is slowing down for many age groups. This is particularly noticeable for infant mortality and for people aged 65 to 74 or above age 75, for whom mortality plateaus are visible. It echoes the increase in mortality for men aged 15 to 34 from 1985 to 1995, which was due to AIDS. Figure 2 also reveals a strong slowdown in the absolute convergence of regional mortality rates for women aged 35 to 64 and for both men and women over age 75. This slowdown results in an increase in relative regional inequalities at these ages. For men above age 75, absolute regional inequalities have increased between 2004 and 2010 and have returned to levels recorded in 2001 when the mortality level was 25% higher. A comparison of the male and female curves demonstrates that male mortality at ages 35 to 64 in 2019 has just reached the female mortality from 1971. Interestingly, men appear to follow a higher path of relative regional inequality than women for ages higher than 15: regional variation in exposure to both occupational hazards and risk-taking behaviour are stronger among men than women.

**Figure 2.**
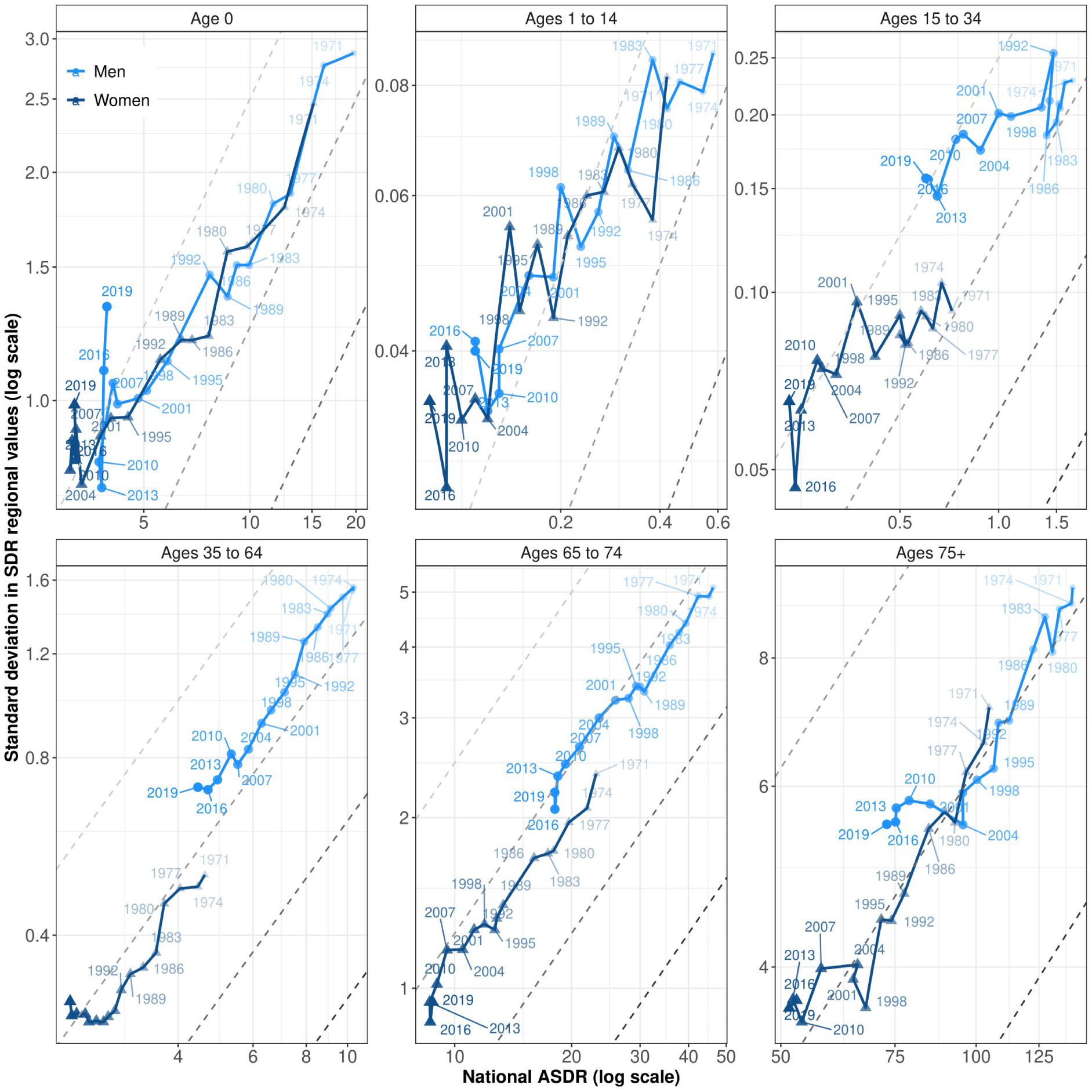
Pathways of national SDRs and regional inequalities in France for six age groups (1970–2019).

Figure 3 presents the same age-specific pathways for Germany. For both men and women in age groups 35 to 64, 65 to 74, and for women aged 15 to 34, the general picture is similar to the one depicted by Figure 1: before 2004, the decline in mortality mirrored an even greater decline in absolute regional inequality, which resulted in a decline in relative regional inequality as well. After 2004, absolute regional inequality stalled while mortality at the national level continued to decline, which resulted in an increase in relative regional inequality. For both men and women in age groups 35 to 64 and 65 to 74, the low decline in mortality in recent years comes along with a worrying increase in absolute regional inequalities. This “Matthew Effect” [34], when national mortality decrease is paralleled by regional inequality increase, contrasts with the strong relationship we globally observe between nationwide mortality and absolute regional inequality. For women aged 35 to 64, this increase is mostly driven by regional differences in smoking behaviour [35]: mortality rates for these ages are stalling or increasing in some regions of central Germany. Finally, the decline in infant mortality between 1992 and 2019 was associated with different phases of convergence (1992–1998; 2007–2019) and divergence (1998–2007).

**Figure 3.**
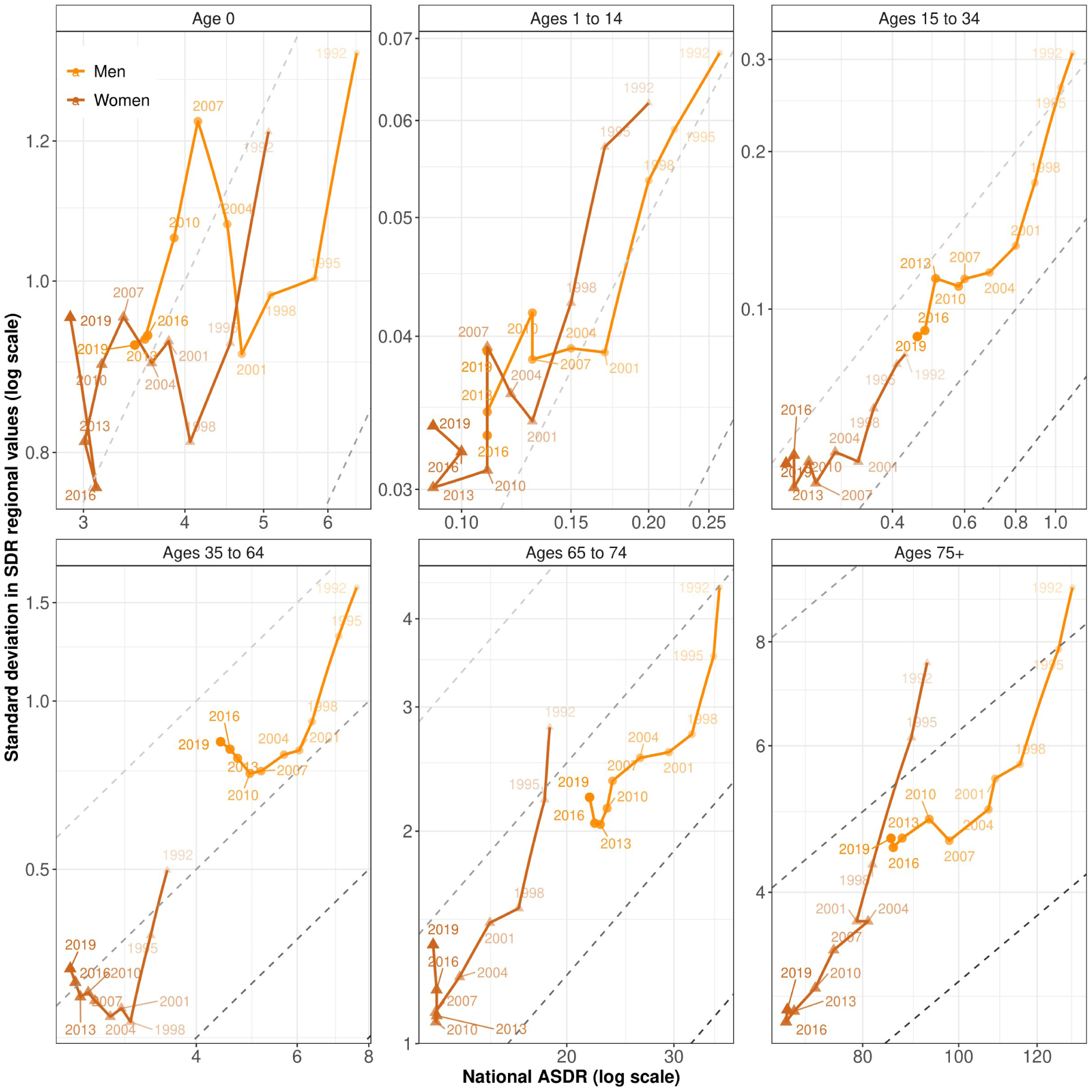
Pathways of national SDRs and regional inequalities in Germany for six age groups (1992–2019).

While Figures 1 to 3 present the global pathways of societal progress in mortality, Figures 4 and 5 emphasize the short-term variations of the two objectives presented in the “Data and methods” section; they illustrate what we call the “*target of societal progress in mortality*” for France and Germany. Each point plots the relative variation in the national SDR (x-axis) and standard deviation (y-axis) between two successive 3-year periods. The point 1995, for example, refers to the relative variation of both national SDR and standard deviation between the 3-year period centered around 1992 and the 3-year period centered around 1995.

**Figure 4.**
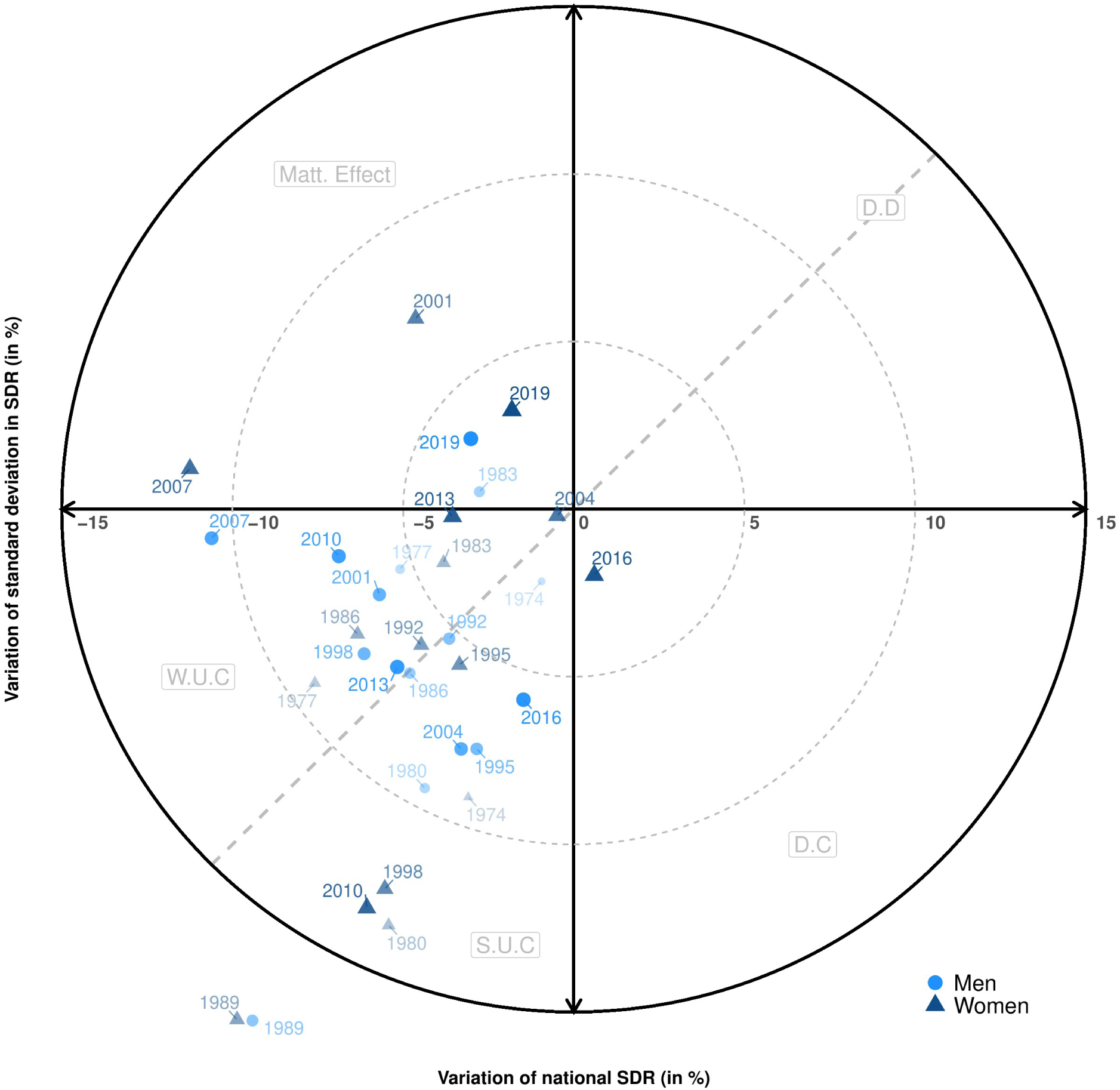
Target of societal progress in mortality in France, 1970–2019

**Figure 5.**
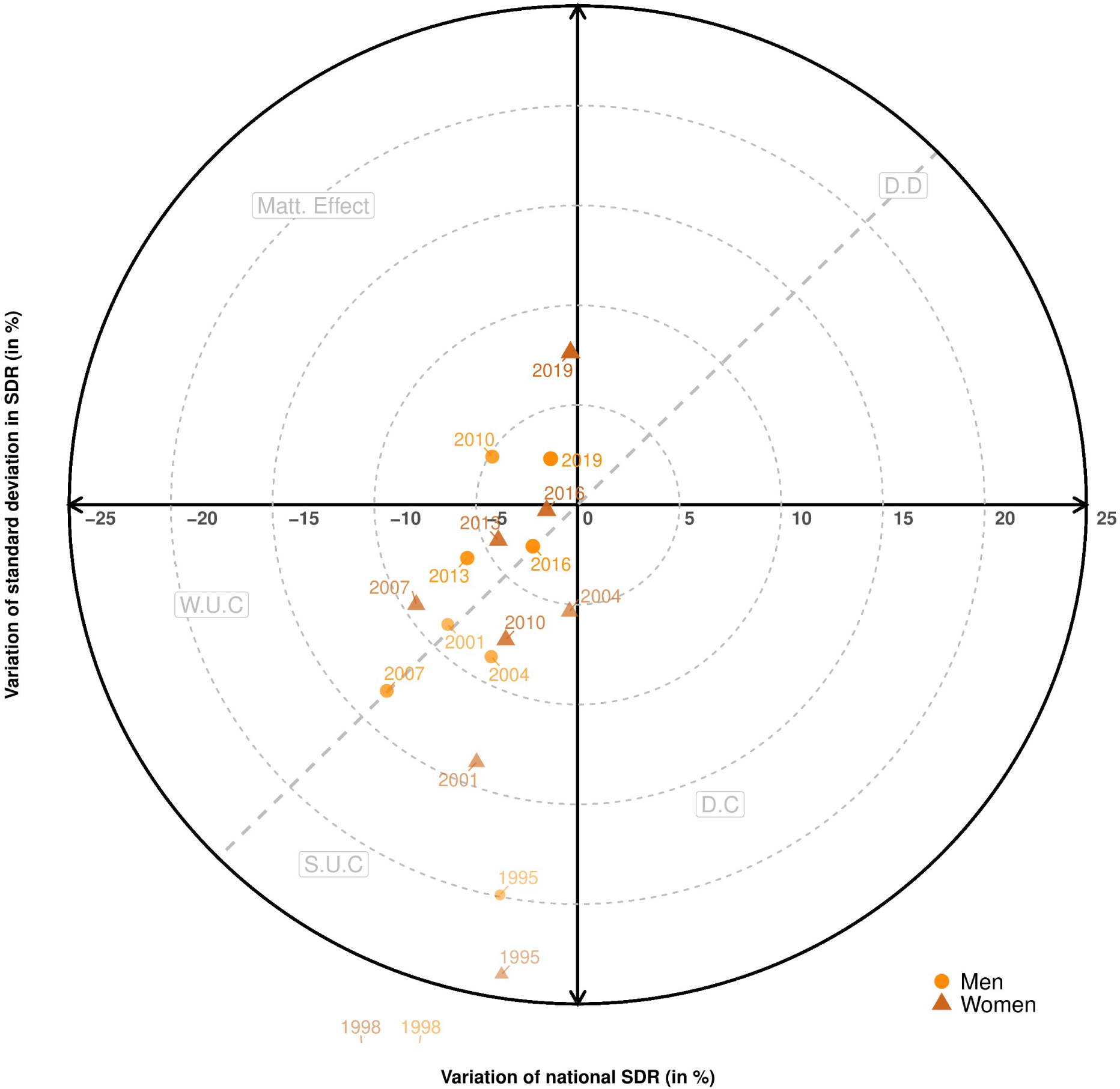
Target of societal progress in mortality in Germany, 1992–2019

The upper left of the target depicts “Matthew Effect” periods [34], when national mortality decreases but regional inequality increases. Health progress benefits the vanguard regions during these periods, and their advantage increases. The upper right of the target depicts periods of “downward divergence” (DD), when both regional inequality and national mortality increases. The lower right of the target depicts “downward convergence” (DC) periods, when regional inequality decreases but national mortality increases. The mortality spike affects the vanguard regions and reduces regional inequality. The lower left of the target depicts “upward convergence” (UC) periods, when both national mortality and absolute inequalities decrease. In this very specific quadrant, we disentangle “strong upward convergence” (SUC) and “weak upward convergence” (WUC) periods. In SUC (resp. WUC) periods, below (resp. above) the dashed line, the decrease in absolute inequality is higher (resp. lower) than the decrease in national mortality, leading to a decrease (resp. increase) in relative inequality.

Figure 4 presents results for France by differentiating men (lighter colours and full circles) and women (darker colours and full triangles). Points are more transparent and smaller the further away they are in time. Overall, most of the points are located in the lower left quadrant of the target, which symbolizes societal progress in mortality: national mortality is decreasing, and so are absolute regional inequalities. Nevertheless, a substantial part of these points are located outside the SUC part of the target, which combines a decline in national mortality, absolute regional inequalities and relative regional inequalities. For women, we observe that absolute regional inequalities decreases are decelerating: leaving aside 2010, triangles are moving to the upper part of the target. Absolute inequalities even reaugment between 1998 and 2001 (+5%), 2004 and 2007 (+1%), and 2016 and 2019 (+3%), Moreover, mortality increased between 2013 and 2016 (+1%) and then decreased weakly between 2016 and 2019 (2%).

Figure 5 presents results for Germany. As revealed by Figure 1, societal progress in mortality was strong in this country during the 1990s. The values reached by the decreasing rate in absolute regional inequalities were particularly high between 1995 and 1998 (outside of the range, between –30 and –40%). Overall, all the points are located in the “strong upward convergence” part of the target before 2007, even if values are moving up. From 2007 onwards, almost all points are located to the left of the dashed line and reveal an increase in relative regional inequalities. Moreover, points are also gradually moving towards the center of the target: improvement rates in global mortality and absolute regional inequalities are close to 0 for the periods 2013–2016 and 2016–2019. Once again, absolute inequalities increased dramatically between 2016 and 2019 for women.

## Conclusion

In this paper, we proposed a new visual approach to assess simultaneously two dimensions of societal progress in longevity by combining (1) global mortality and (2) inequality measures. Due to the mathematical properties of the indicators used, one can simultaneously grasp the evolution of absolute and relative inequalities.

Before discussing the implications of our visual representation, we have to acknowledge its few limitations. One drawback of our visualisation tool is that it requires medium– or long-term data to clearly represent the pathways of societal progress in each population and make relevant comparisons between populations whose mortality rates are not at the same level nowadays. Moreover, comparisons must be made between homogeneously stratified populations: in our application, a roughly similar number of regions was crucial to interpret the differences between France and Germany. However, this is less of an issue with other stratification approaches such as by level of education or income, which could also be used. For such approaches, it is usually easier to derive cross-country comparable stratifications with a similar number of categories.

In our case study, which focused on regional mortality inequalities, we demonstrated how our proposed visual approach is able to depict the evolution of societal progress over time. In both France and Germany, the pace of societal progress in mortality has decreased, and it has remained at the almost nil level since 2013 for both men and women. More alarmingly, for both men and women aged 35 to 74 in Germany, the low decline in mortality in recent years is accompanied by increasing absolute regional inequalities in mortality. This contrasts with the strong relationship we globally observe between national mortality and absolute regional inequality. More precisely, the increase in regional inequality for women aged 65–74 occurs with unchanged mortality, which could indicate that some regions are experiencing an increase in mortality.

We also show how this representation allows better comparisons by sex or across countries since the differences in global mortality observed partly explain the greater or lesser absolute regional inequality. In particular, we revealed that the sex-specific pathways of societal progress are better in Germany than in France since absolute regional inequalities are lower for all values of death rates except during the early 1990s. Moreover, the pathways are also more favourable for women than for men in both countries.

In summary, thanks to its flexibility and low computational cost, we envisage a wider use of our framework for monitoring societal progress in mortality at the international level. Given this purpose, we provide R routines (see Online Supplementary Material) to replicate our graphical representations in other countries and for different population strata.

## Data Availability

All data produced are available online at:
https://osf.io/h68wz/?view_only=47353f6f4b2e41cab3c761246e59d615

## Online Supplementary Appendix A

**Figure A1:**
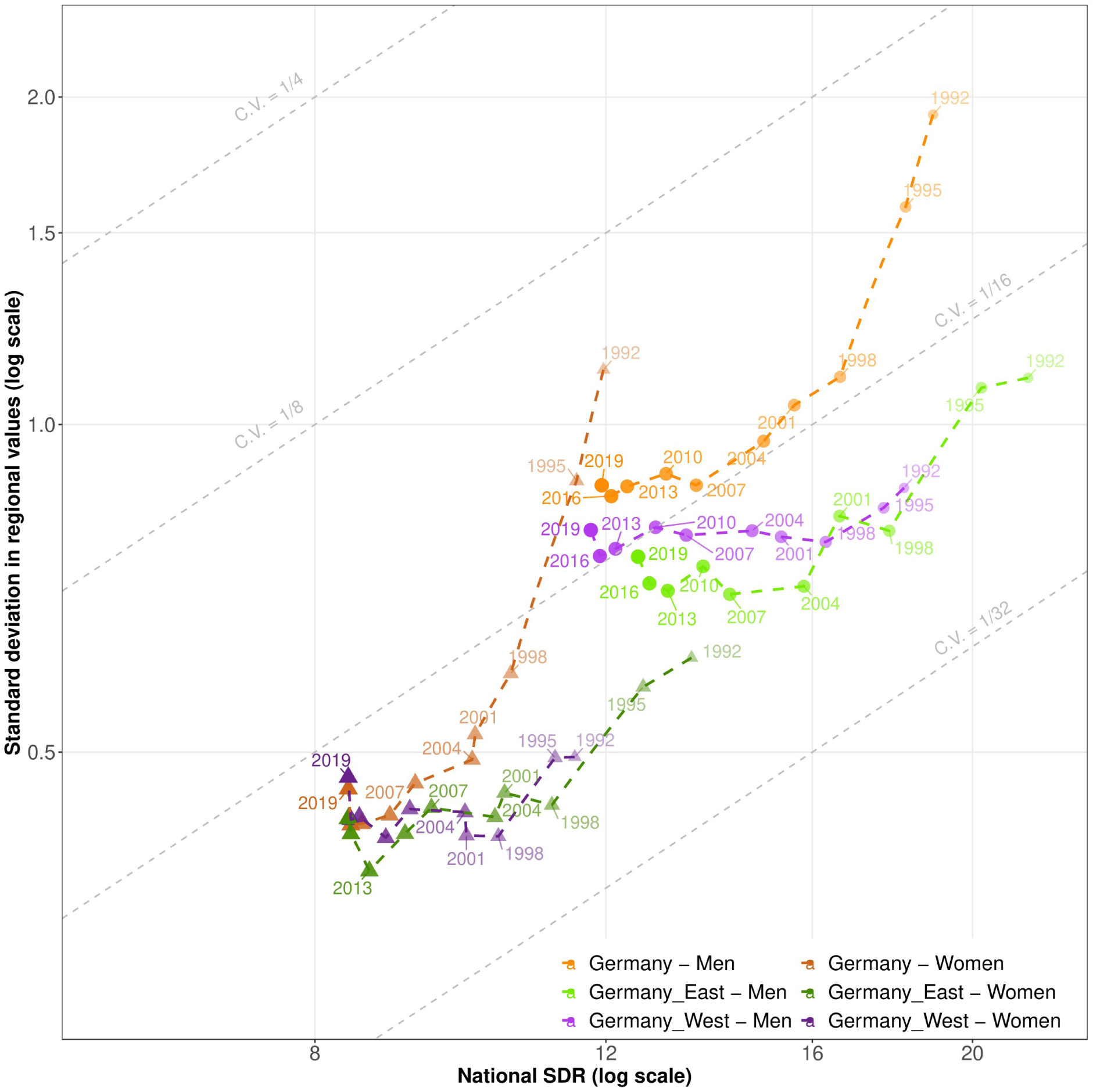
Pathways of overall SDRs and regional inequalities in western, eastern and total Germany, 1992–2020.

## Online Supplementary Appendix B

Data and R code to replicate our figures are available at: https://osf.io/h68wz/?view_only=47353f6f4b2e41cab3c761246e59d615

Please read first “Online Supplementary Appendix.pdf”.

